# Head-to head comparison of anterior nares and nasopharyngeal swabs for SARS-CoV-2 antigen detection in a community drive-through test centre in the UK

**DOI:** 10.1101/2022.09.06.22279637

**Authors:** Rachel L Byrne, Ghaith Aljayyoussi, Konstantina Kontogianni, Karina Clerkin, Mathew McIntyre, Jahanara Wardale, Christopher T Williams, CONDOR steering group, Richard Body, Emily R Adams, Margaretha de Vos, Camille Escadafal, Ana I Cubas Atienzar

## Abstract

**Objective:** To conduct a head-to-head diagnostic accuracy evaluation of professionally taken anterior nares (AN) and nasopharyngeal (NP) swabs for SARS-CoV-2 antigen detection using rapid diagnostic tests (Ag-RDT).

**Methods:** NP swabs for SARS-CoV-2 reverse transcription quantitative polymerase chain reaction (RT-qPCR) testing and paired AN and NP swabs for the antigen detection were collected from symptomatic participants enrolled at a community drive-through COVID-19 test centre in Liverpool. Two Ag-RDT brands were evaluated: Sure-Status (PMC, India) and Biocredit (RapiGEN, South Korea). The visual read out of the Ag-RDT test band was quantitative scored and the 50% and 95% limit of detection (LoD) of both Ag-RDT brands using AN and NP swabs was calculated using a probabilistic logistic regression model.

**Results:** A total of 604 participants were recruited of which 241 (40.3%) were SARS-CoV-2 positive by RT-qPCR. Sensitivity and specificity of AN swabs was equivalent to the obtained with NP swabs: 83.2% (75.2-89.4%) and 98.8% (96.5-99.6%) utilising NP swabs and 84.0% (76.2-90.1%) and 99.2% (97.0-99.8%) with AN swabs for Sure-Status and; 81.2% (73.1-87.7%) and 99.0% (94.7-86.5%) with NP swabs and 79.5% (71.3-86.3%) and 100% (96.5-100%) with AN swabs for Biocredit. The agreement of the AN and NP swabs was high for both brands with an inter-rater relatability (κ) of 0.918 and 0.833 for Sure-Status and Biocredit, respectively. The overall 50% LoD and 95% LoD was 0.9-2.4 × 10^4^ and 3.0-3.2 × 10^8^ RNA copies/mL for NP swabs and 0.3-1.1 × 10^5^ and 0.7-7.9 × 10^7^ RNA copies/mL and for AN swabs with no significant difference on LoD for any of the swabs types or test brands. Quantitative read-out of test line intensity was more often higher when using NP swabs with significantly higher scores for both Ag-RDT brands.

**Conclusions:** the diagnostic accuracy of the two SARS-CoV-2 Ag-RDTs brands evaluated in this study was equivalent using AN swabs than NP swabs. However, test line intensity was lower when using AN swabs which could influence negatively the interpretation of the Ag-RDT results for lay users. Studies on Ag-RDT self-interpretation using AN and NP swabs are needed to ensure accurate test use in the wider community.

## Introduction

To meet the immense diagnostic demand of the COVID-19 pandemic, the development of rapid diagnostic tests for the detection of SARS-CoV-2 antigens (Ag-RDTs) became a priority [1]. Nasopharyngeal (NP) swabs are considered the standard of care for SARS-CoV-2 detection[2] and thus the majority of Ag-RDT kits are developed for NP swabs exclusively [1]. However, the use of anterior nasal (AN) swabs has been increasing as a less invasive alternative to promote access to testing in the community and facilitate mass testing programmes particularly in the UK [3].

For Ag-RDTs, studies on Ag-RDTs comparing sensitivity on AN swabs and NP swabs are very limited, with only two reported studies performed on the same Ag-RDT brand, Standard-Q (SD Biosensor, Inc., Korea), one study on professional taken swabs [6] and another in self-taken [7]. Sensitivity obtained with AN swabs was comparable (although 3% to 5% lower) than with NP swabs sensitivity but neither of the swab types fulfilled WHO target product profile (TPP) standards in any of the two studies [8]. AN swabs are considered accurate and clinically acceptable alternatives to NP swabs in outpatient settings for SARS-CoV-2 reverse transcription polymerase chain reaction (RT-PCR) testing [4]. However, an in depth metanalysis on SARS-CoV-2 RT-PCR testing found that anterior nares specimens were 12%-18% less sensitive than NP swabs [5].

The aim of this study was to perform a head-to-head evaluation on two World Health Organisation (WHO) approved or under assessment for Emergency Use listing (WHO-EUL) SARS-CoV-2 Ag-RDT brands marketed for AN and NP swabs: Sure-Status COVID-19 Antigen Card Test (Premier Medical Corporation, India) and Biocredit COVID-19 Antigen Test (RapiGEN, South Korea) respectively. This study is of particular interest in the UK as the use of home Ag-RDTs on AN swabs has been integral to combatting the spread of COVID-19 during the pandemic [3], as on the 1^st^ of April 2022 free national RT-PCR COVID-19 testing was suspended, with the purchase of Ag-RDTs using AN-swabs online or in pharmacies the only approach to access COVID-19 testing in a non-clinical setting.

## Methods

### Clinical evaluation

This was a prospective evaluation of consecutive participants enrolled at a community National Health Service (NHS) drive-through COVID-19 test centre located at the Liverpool John Lennon Airport. Two Ag-RDT brands were evaluated; Sure-Status COVID-19 Antigen Card Test (Premier Medical Corporation India) and Biocredit COVID-19 Antigen Test (RapiGEN, South Korea) referred as Sure-Status and Biocredit thereafter. The study progressed until at least 100 Ag-RDT positives using AN swabs in line with WHOs requirements for evaluation of alternative sample type [10].

All adults over the age of 18 who attended the drive-through test centre with symptoms of COVID-19 were asked to participate in the study. The symptoms included fever, cough, shortness of breath, tight chest, chest pain, runny nose, sore throat, anosmia, ageusia, headache, vomiting, abdominal pain, diarrhoea, confusion, rush, or tiredness. Participants were recruited under the Facilitating Accelerated COVID-19 Diagnostics (FALCON) study using verbal consent. Ethical approval was obtained from the National Research Ethics Service and the Health Research Authority (IRAS ID:28422, clinical trial ID: NCT04408170).

Swabs were collected following the same process with the NP swab collected first in one nostril and placed in Universal Transport Media (UTM) (Copan Diagnostics Inc, Italy) for the reference RT-qPCR test. This was followed by the collection of two swabs to evaluate the Ag-RDTs, first an NP swab in the other nostril and finally a AN swab in both nostrils following the manufacturer’s instructions for use (IFU). Samples were given a unique identification code and transported within cooler bags to the Liverpool School of Tropical Medicine (LSTM) where samples were processed in category level 3 (CL3) containment laboratory upon arrival.

Sure-Status and Biocredit Ag-RDTs were carried out following their instructions for use (IFU). The protocol for both Ag-RDT was the same when using AN and NP swabs. Results were read by two operators, blinded to one another and if a discrepant result occurred, a third operator acted as a tiebreaker. The visual read out of the Ag-RDT test band was scored on a quantitative scale from 1 (weak positive) - 10 (strong positive). Ag-RDT results were classified as invalid when the control line was absent.

RNA was extracted using the QIAamp® 96 Virus QIAcube® HT kit (Qiagen, Germany) on the QIAcube® (Qiagen, Germany) and screened using TaqPath COVID-19 (ThermoFisher, UK) on the QuantStudio 5™ thermocycler (ThermoFisher, UK), an internal extraction control was incorporated before the lysis stage, as recommended by the manufacturer. SARS-CoV-2 RT-qPCR result was considered (1) positive if any two of the three SARS-CoV-2 target genes (N gene, ORF1ab and S gene) amplified with cycle threshold (Ct) ≤ 40, (2) indeterminate if only one SARS-CoV-2 gene amplified and (3) negative if the internal extraction control amplified and the SARS-CoV-2 target genes did not. Samples with invalid RT-qPCR results (no amplification of the internal extraction control) were re-extracted and re-run once. Viral loads in UTM swabs were measured with a ten-fold serial dilution standard curve of quantified specific in vitro-transcribed RNA using five replicates for each standard curve point [11].

### Statistical Analysis

Sensitivity, specificity, positive predicted value (PPV) and negative predictive values (NPV) were calculated with 95% confidence intervals (CIs) by comparing the Ag-RDT results to the RT-qPCR, as the reference standard. Sub-analyses of diagnostic performance were performed by swab type (AN and NP), Ct-value ranges, onset of symptoms and vaccination status using nonparametric statistics. The level of agreement between AN and NP swabs was determined using Cohen’s kappa (κ) [10]. The correlation between test line intensity and viral loads were measured by Person correlation, coefficient (r_P_) [12] and to further analyse Ag-RDT sensitivities, we used logistic regression, with RNA copy numbers of the RT-qPCR NP swab and swab type (AN and NP) as independent variables and test outcomes as the dependent variable, yielding detection probabilities for each viral load level. Statistical analyses were performed using SPSS V.28.0, Epi Info V3.01 and R scripts. Statistical significance was set at *P* < 0.05.

## Results

### Participant demographics

A total of 604 participants were recruited for this study, 372 recruited between August and October 2021 were enrolled for the Sure-Status Ag-RDT evaluation and 232 recruited between December 2021 and March 2022 were enrolled for the Biocredit Ag-RDT evaluation. Details of the demographics of the population of study are found in Table 1. Our study population had a mean age of 43 years (range 18-81, interquartile range [IQR] 33.0-50.0), 348 (58%) were female and 566 were British (94%), with the remaining 36 participants being of other ethnic groups (n = 14), white background (n = 9), Asian (n = 8), mix white and black backgrounds (n = 2) and Arab (n = 1). Three hundred and fourteen participants of the 372 enrolled for the Sure-Status evaluation (84.4%) and 217 participants of the 232 recruited for Biocredit (93.5%) received complete SARS-CoV-2 vaccination (2 doses). Additionally, 143 of the participants enrolled from December 2021 (61.6%) for the Biocredit evaluation received a third dose as part of the UK booster roll out [13]. All participants were symptomatic with a median onset of symptoms of 2 days (IQR 1-3). The most common symptoms were cough (387, 64.3%), sore throat (232, 38.5%), headache (203, 33.7%), fever (160, 26.6%), body aches (80, 13.3%) and runny nose (80, 13.3%) (Table 1).

**Table 1.** Demographics of the population of study for Sure-Status and Biocredit cohorts

|  | Sure-Status | Biocredit | All |
| --- | --- | --- | --- |
| Total | 372 | 232 | 604 |
| Age [mean (min-max), IQR] | 43 (18-81), 33-53 | 43 (18-78), 33-51 | 43 (18-81), 33-52 |
| Gender [%F, (n/N)] IQR] | 57%, (211/372) | 59%, (137/232) | 58%, (348/602) |
| Triple vaccinated (n, %) | NA* | 143 (61.6%) | 143 (23.8%) |
| Double vaccinated (n, %) | 314 (84.4%) | 74 (40%) | 388 (64.4%) |
| Partially vaccinated (n, %) | 29 (7.8%) | 4 (1.7%) | 33 (5.5%) |
| Not vaccinated (n, %) | 27 (7.3%) | 10 (4.3%) | 37 (6.2%) |
| Vaccination not disclosed (n, %) | 2 (0.5) | 1 (0.3%) | 3 (0.5%) |
| Days symptoms onset [median (IQR); N] | 2 (1-3), 371 | 2 (1-3), 232 | 2 (1-3), 601 |
| Days 0-3 (n, %) | 304, 81.7% | 186, 80.2% | 490, 81.1% |
| Days 4-7 (n, %) | 56, 15.1% | 41, 17.7% | 97, 16.1% |
| Days 8+ (n, %) | 10, 2.7% | 5, 2.2% | 15, 2.5% |
| RT-qPCR SARS-CoV-2 Positivity [%, (n/N)] | 32.1%, (119/371) | 53.7%, (122/227) | 40.3% (241/598) |
| Symptom [total n (%), in RT-qPCR positive n (%)] |  |  |  |
| Cough | 248 (66.7%), 73 (61.3%) | 139 (60.0%), 71 (58.2%) | 387 (64.3%), 144 (60.0%) |
| Sore throat | 129 (34.7%), 34 (28.6%) | 103 (44.4%), 56 (45.9%) | 232 (38.5%), 90 (37.4%) |
| Headache | 123 (33.1%), 57 (47.9%) | 80 (34.5%), 45 (36.9%) | 203 (33.7%), 102 (42.3%) |
| Fever | 106 (28.5%), 30 (25.2%) | 54 (23.3%), 28 (22.9%) | 160 (26.6%), 58 (24.1%) |
| Body aches | 41 (11.0%), 21 (17.7%) | 39 (16.8%), 29 (23.8%) | 80 (13.3%), 51 (21.2%) |
| Runny nose | 39 (13.2%), 20 (16.8%) | 41 (17.7%), 31 (25.4%) | 80 (13.3%), 51 (21.2%) |
| Loss taste | 48 (12.9%), 19 (16.0%) | 19 (8.2%), 10 (8.2%) | 67 (11.1%), 29 (12.0%) |
| Loss smell | 29 (7.8%), 9 (7.6%) | 14 (6.0%), 7 (5.7%) | 43 (7.1%), 16 (6.6%) |
| Chest pain | 18 (4.8%), 7 (5.9%) | 12 (5.2%), 8 (6.6%) | 30 (5.0%), 15 (6.2%) |
| Fatigue | 13 (3.5%), 4 (3.4) | 17 (7.3%), 10 (8.2%) | 30 (5.0%), 14 (5.8%) |
| Shortness of breath/tight chest | 13 (3.5%), 3 (2.5%) | 9 (3.9%), 5 (4.1%) | 22 (3.6%), 15 (6.2%) |
| Vomiting | 11 (3%), 5 (4.2%) | 2 (8.6%), 2 (1.6%) | 13 (2.2%), 7 (2.9%) |
| Diarrhoea | 9 (2.4%), 3 (2.5%) | 3 (13%), 3 (2.5%) | 12 (2.0%), 6 (2.5%) |
| Abdominal pain | 6 (1.6%), 3 (2.5%) | 1 (0.4%), 1 (0.8%) | 7 (1.2%), 4 (1.7%) |
| Rash | 3 (0.8%), 0 (0.0%) | 1 (0.4%), 1 (0.8%) | 4 (0.6%), 1 (0.4%) |
| Confusion | 1 (0.3%), 0 (0.0%) | 0 (0%) | 1 (0.2%), 0 (0%) |
| Other | 159 (42.7%), 68 (57.4%) | 134 (57.8%), 85 (69.7%) | 293 (48.7%), 153 (63.5%) |

\*Participants were enrolled before booster rolled out in the UK

Overall, 241 participants (40.3%, CI95% 36.3-44.4%) were SARS-CoV-2 positive by RT-qPCR, 6 had indeterminate RT-qPCR results and the remaining 355 were negative. Participants with indeterminate RT-qPCR results were excluded from further analysis.

RT-qPCR positivity was significantly higher (p<0.05) among the participants enrolled for the Biocredit evaluation cohort (53.7%, CI95% 47-60.4%) during December 2021 and March 2022 which coincided with the Omicron wave in the UK [14] than among the participants enrolled between August and October 2021 (32.1%, CI95% 27.4-37.1%) when Delta was the dominant SARS-CoV-2 variant.

### Diagnostic evaluation

#### Sure Status

The overall sensitivity and specificity for the Sure-Status Ag-RDT compared to RT-qPCR was 83.2% (CI95% 75.2-89.4%) and 98.8% (CI95% 96.5-99.6%) utilising NP swabs and 84.0% (CI95% 76.2-90.1%) and 99.2% (CI95% 97.0-99.8%) with AN swabs. For individuals with Cts < 25, the sensitivity was 91.8% (CI95% 84.5-96.4%) and 93.8% (CI95% 87.2-97.7%) for NP and AN-swabs respectively. Seven Ag-RDTs gave invalid results, one NP swab (0.03%) sample and six AN swab samples (1.6%). Participants with invalid Ag-RDTs results were excluded from further analysis. Four SARS-CoV-2 positive cases were detected by NP only (3.4%) and six cases were detected by AN only (5.0%) but this discrepancy on sensitivity between swab types was not significant (*P* = 0.43). The percentage of agreement of NP and AN swab using Sure-Status was 96.7% (95% CI 94.3-98.3%) and inter-rater reliability was almost perfect (κ = 0.918). Inter-rater reliability was strong for both NP (κ = 0.871) and AN (κ = 0.852) swabs when compared to RT-qPCR.

#### Biocredit

For the Biocredit Ag-RDT the sensitivity and specificity were 81.2% (CI95%73.1-87.7%) and 99.0% (CI95%94.7-86.5%) with NP swabs and 79.5% (CI95%71.3-86.3%) and 100% (CI95%96.5-100%) with AN sampling compared to RT-qPCR. Sensitivity was 92.2% (CI95%84.6-96.8%) and 95.5% (CI95%89.0-98.8%) using NP and AN swabs among participants with Ct < 25. Ten SARS-CoV-2 positive cases were detected solely by NP (8.2%) and eight cases were detected only by AN (6.6%) but no significance on sensitivity was observed between NP and AN swabs for this brand of Ag-RDTs either (*P* = 0.43). The percentage of agreement of NP and AN swab for Biocredit was 91.6% (95% CI 87.2-94.9%) and inter-rater reliability was strong (κ = 0.833). Inter-rater reliability was moderate for both NP (κ = 0.790) and AN (κ = 0.782) sampling compared to RT-qPCR. Diagnostic accuracy for both Sure-Status and Biocredit is displayed in Table 2.

**Table 2.** Clinical sensitivity and specificity of Sure-Status and Biocredit using NP and Nasal Swab

| All Ct values | TP | FP | TN | FN | Sensitivity | Specificity | NPV | PPV |
| --- | --- | --- | --- | --- | --- | --- | --- | --- |
| Sure-Status | 200 | 5 | 494 | 38 | 83.9 (78.3-88.1) | 99.0 (97.7-99.7) | 92.7(90.5-94.4) | 97.6 (94.3-98.9) |
| NP swab | 99 | 2 | 249 | 20 | 83.2 (75.2-89.4) | 98.8 (96.5-99.6) | 91.7 (88.4-94.2) | 97.1 (91.44-99.03) |
| AN swab | 101 | 2 | 245 | 18 | 84.0 (76.2-90.1) | 99.2 (97.0-99.8) | 92.8 (89.5-95.1) | 98.0 (92.62-99.5) |
| Biocredit | 196 | 1 | 209 | 48 | 80.3 (74.8-85.1) | 99.1 (95.0-99.9) | 81.3(77.2-84.9) | 99.5 (96.5-100) |
| NP swab | 99 | 1 | 104 | 23 | 81.2 (73.1-87.7) | 99.0 (94.7-86.5) | 81.9 (73.99-85.2) | 99.0 (93.4-99.9) |
| AN swab | 97 | 0 | 105 | 25 | 79.5 (71.3-86.3) | 100 (96.5-100) | 80.8 (74.8-85.6) | 100 (100-100) |
| All NP | 198 | 4 | 353 | 43 | 82.2 (76.7-86.8) | 98.9 (97.2-99.7) | 89.1 (86.2-91.5) | 98.0 (94.9-99.2) |
| All AN | 198 | 2 | 350 | 43 | 82.2 (76.7-86.8) | 99.4 (97.9-99.9) | 89.1 (86.2-91.5) | 99.0 (96.1-99.8) |

#### Head to head comparison of Sure Status and Biocredit

rWe report nosignificant difference in the diagnostic accuracy among participants with symptoms irrespective of days since onset, or vaccination status for all Ag-RDTs and swabbing combination (all *P* values > 0.05). Both Biocredit and Sure-Status Ag-RDTs using both swab types had better sensitivities on detecting SARS-CoV-2 antigens on individuals with Ct values < 25 than >30 (*P* = 0.029 in NP and *P* = 0.047 in AN for Sure-Status and *P* = 0.018 and *P* = 0.001 for Biocredit).

The RNA copy numbers per mL (RNA copies/mL) of RT-PCR NP swabs was calculated and statistically higher viral loads were obtained for the Sure-Status cohort than Biocredit (Fig 1) measured by Kruskal–Wallis (P= 0.006). We determined the 50% and 95% limits of detection (LoD) for both Ag-RDT and swab types based on a logistic regression model (Fig 2). For Sure-Status, the RNA copies/mL for 50% LoD and 95% LoD were 2.4 × 10^4^ and 3.16 × 10^8^ for NP specimen and 3.4 × 10^4^ and 7.94 × 10^7^ for AN swabs. For Biocredit, the RNA copies/mL for LoD50 and LoD95 were 9.12 × 10^3^ and 3.02 × 10^8^ for NP specimen and 1.12 × 10^5^ and 6.76 × 10^6^ for AN swabs. Although the LoD95 was better for AN swabs for both Ag-RDT brands (3.98 for Sure-Status and 44.67 for Biocredit), there was no statistical difference on LODs neither by swab type and Ag-RDT brand (all *P* values > 0.05).

**Figure 1.**
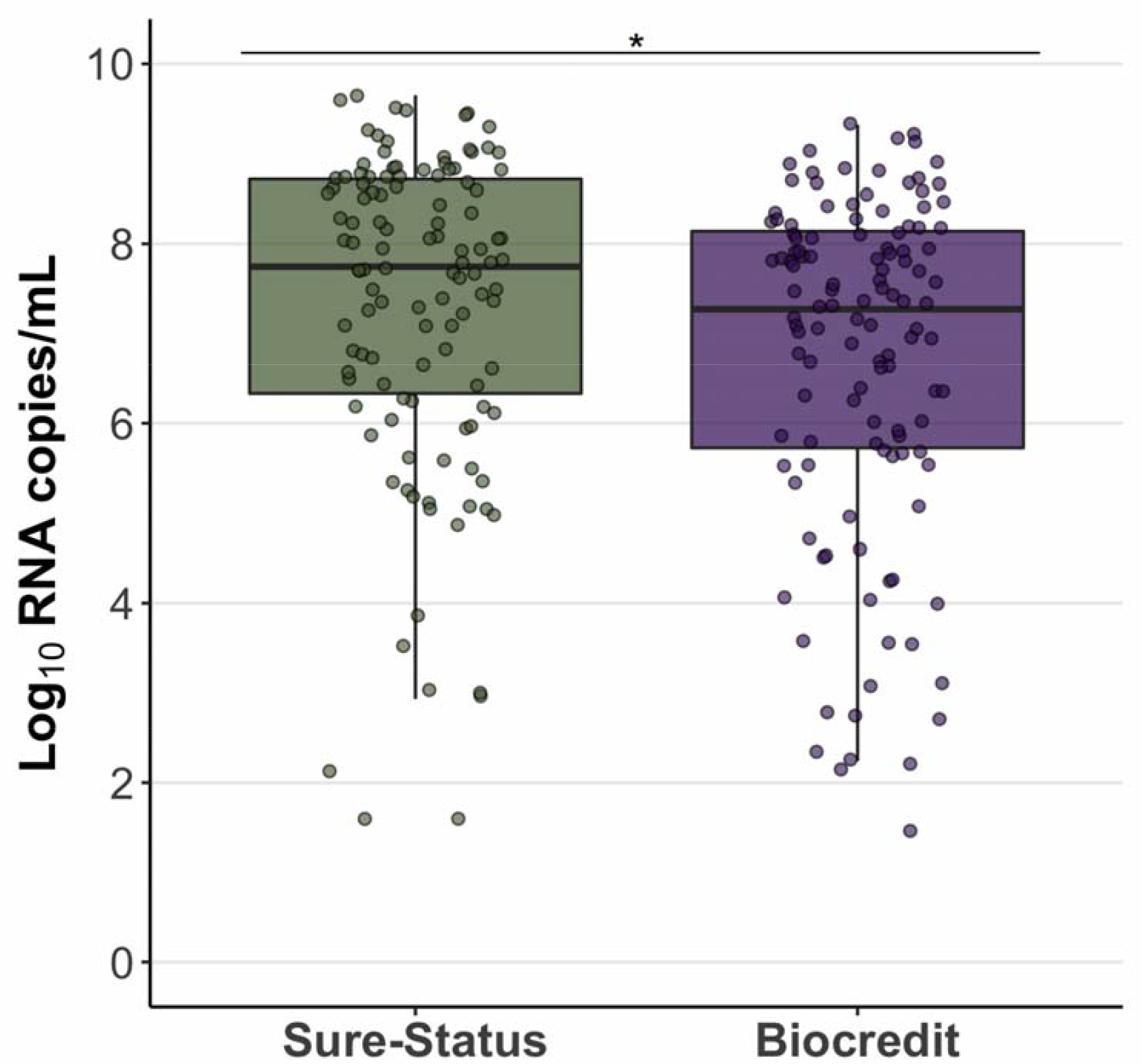
Boxplot of the SARS-CoV-2 viral load distribution of the RT-qPCR NP swabs used as reference standard for the participants enrolled for Sure-Status and Biocredit Ag-RDT evaluation. The whiskers show the maximum and minimum values and the vertical line the median. Asterisks indicate statistical significance between AN and NP swab types.

**Figure 2.**
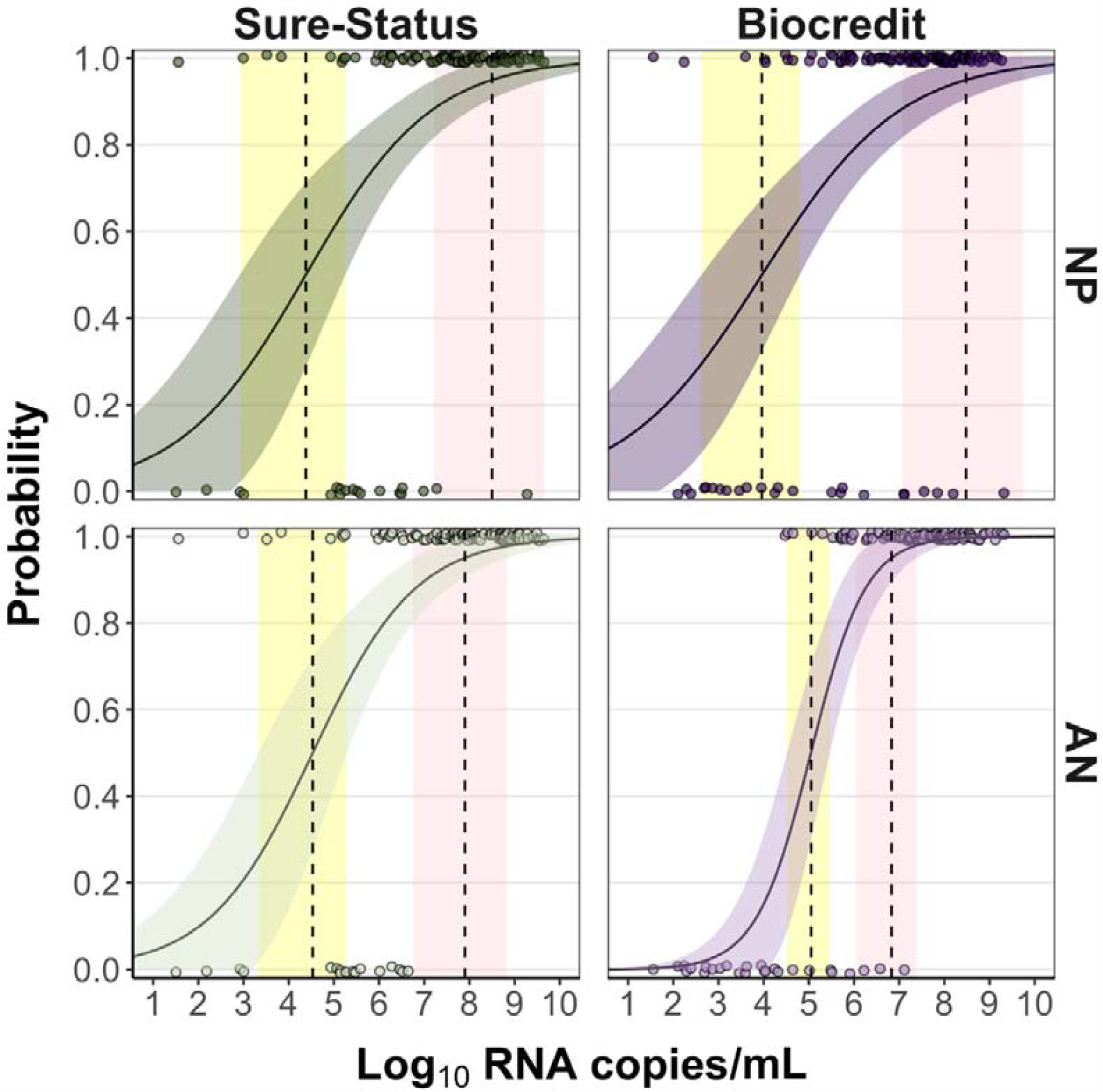
Limit of detection analyses of upper-respiratory samples positive by RT-qPCR for Sure-Status and Biocredit using AN and NP swabs. The log10 RNA copies on the x axis were plotted against a positive (1.0) or negative (0.0) Ag-RDT result on the y axis. Green (Sure-Status) and purple (Biocredit) curves show logistic regressions of the viral load on the Ag-RDT result; vertical dashed lines indicate log10 RNA copies subjected to the test at which 50% and 95% LoD of the samples are expected positive based on the regression results. No significant differences were observed.

**Figure 3.**
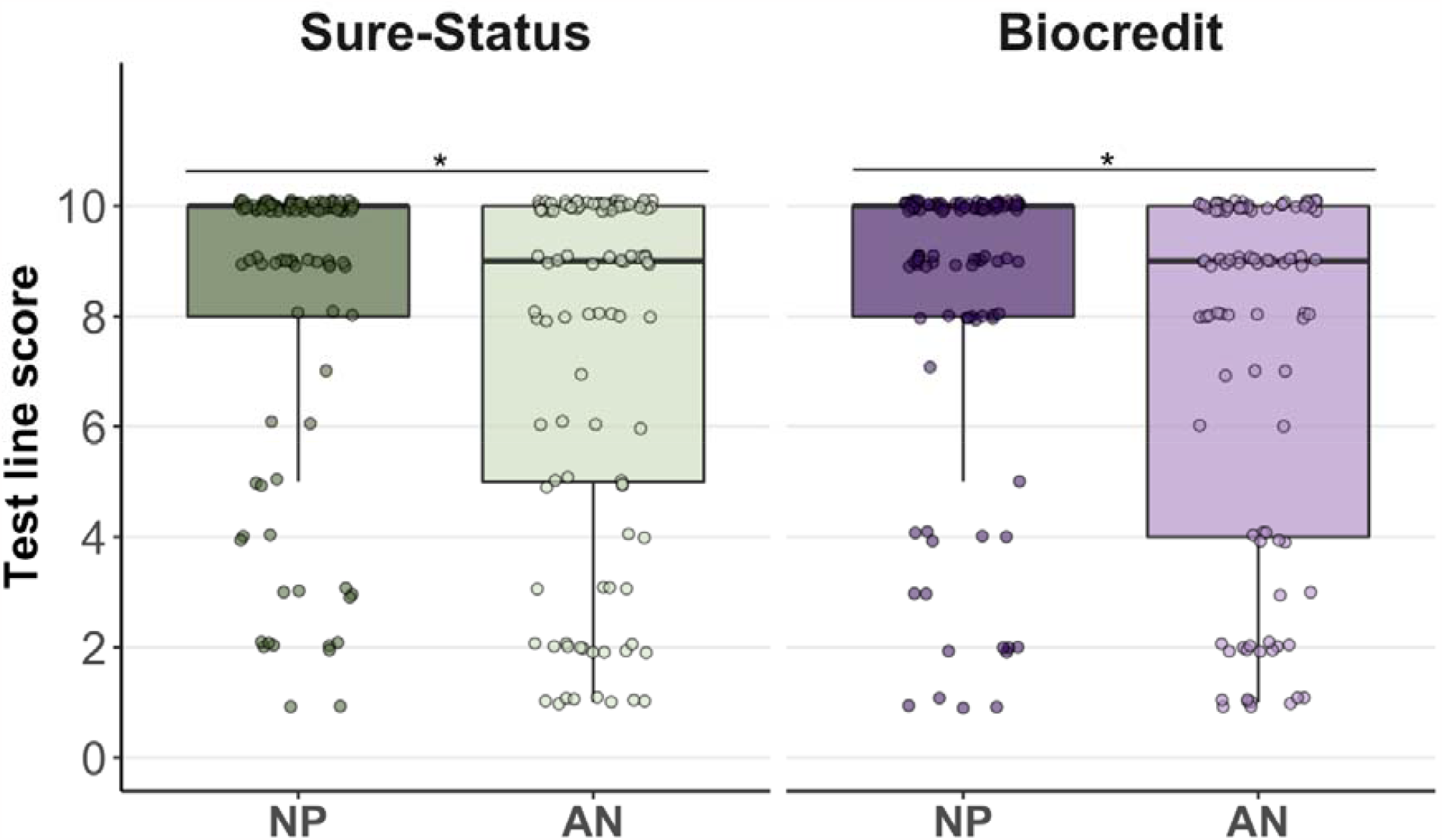
Boxplot of the scores of the test lines for both Ag-RDT Sure-Status and Biocredit using AN and NP swabs. The whiskers show the maximum and minimum values and the vertical line the median. Asterisks indicate statistical significance between AN and NP swab types.

#### Quantitative read-out analysis

Quantitative read-out in paired positive AN and NP was more often higher for the NP (40 instances higher on NP and four higher on AN in Sure-Status; and 35 instances higher on NP and 12 higher on AN in Biocredit) and gave significantly higher scores for both Ag-RDT, Sure-Status (*P* = 0.007) and Biocredit (*P* = 0.013) (Figure 1) measured by Kruskal–Wallis. Additionally, test lines scores were analysed by RNA copies/mL and these had a positive correlation. For Biocredit, strong correlation using AN swabs (r_P_ = 0.727) but moderate using NP swabs (r_P_ = 0.591). For Sure-Status, both swab types had a moderate correlation to viral loads (NP swab r_P_ = 0.614 and AN swab r_P_ = 0.661).

## DISCUSSION

This is the first diagnostic clinical evaluation of Sure-Status Ag-RDT and results have shown a satisfactory performance for both AN and NP swabs fulfilling the sensitivity (≥80%) and specificity (≥97%) outlined in the target product profile (TPP) WHO standards [8]. For Biocredit Ag-RDT, there are five studies to date that have evaluated the performance of NP swabs reporting varied sensitivities from 52% to 85% [15]. In this study we report a higher sensitivity (81.2%, CI95%73.1-87.7%) and specificity (99.0%, CI95%94.7-86.5%) of the Biocredit Ag-RDT fulfilled the WHO standards using NP swab but underperformed in the sensitivity (79.5%, CI95% 71.3-86.3%) criteria when using AN.

Results presented here demonstrate that AN swabs are equivalent to NP swabs for SARS-CoV-2 Ag-RDT testing giving comparable sensitivities, 50% LoD and 95% LoD for both Ag-RDTs brands evaluated here. Our results supports previous findings where AN and NP swabs were compared for the Ag-RDT Standard-Q (SD Biosensor, Inc., Korea) in Lesotho [6], but we reported a higher sensitivity compared to the 67.3% and 70.2% for AN and NP swabs previously described [6]. Studies on RT-qPCR have found lower sensitivity using AN swabs compared to NP swabs consistently [5]. However, the difference in sensitivity was only significant for patients with viral loads < 10^3^ copies/mL [16] and this threshold is not relevant to Ag-RDTs of which the limit of detection ranges between 10^4^-10^8^ RNA copies/mL in swabs [11].

Quantitative assessment of the test line scores showed that test line intensity was significantly higher on NP swabs than AN swabs. The line intensity is an important component of home testing as studies have shown fainter lines are more difficult to interpret for a lay person, likely due to lower signal intensity [17]. In an user experience home based study, 77.1% of the cases that the participants interpreted wrongly as negative being positive, were weak and moderate positives while only 22.9% were strong positives [17]. The significantly lower intensity of the AN swab compared to NP swab is likely attributed to the differences of SARS-CoV-2 viral loads in the respiratory tract. Studies have found lower viral loads on AN swabs compared to NP swabs [16]. Statistical analysis supported this hypothesis where a positive correlation between viral loads and Ag-RDT test line scores was shown. Further implementation studies on Ag-RDT test results interpretation by patients or within a home testing setting are urgently needed to drive self-testing to scale.

This study has several strengths, the use of standardised sampling methods, independent blinded readers, robust statistical analysis, quantitative assessment of Ag-RDT test line results and the evaluation of one approved WHO-EUL Ag-RDT test brands (Sure-Status) and under review (Biocredit). Qualifying it to have high global public health relevance [18].

The main limitation of this study is that the AN swabs were always taken last. The order of sample collection could have negatively biased the results obtained for AN swabs caused by a possible sample depletion. However, in the two studies that compared Ag-RDT using AN swabs, the AN swab was collected first and our reported sensitivity and specificity for AN swabs are greater than the previous studies [6,7]. Further, studies on RT-qPCR found lower sensitivity using AN swabs compared to NP swabs [5], even when AN swabs were collected first [15,18,19]. Thereby it is unlikely that the order of the swabs impacted sample availability for AN and NP sampling.

In conclusion, this study demonstrates the sensitivity of two SARS-CoV-2 Ag-RDTs using AN-sampling are comparable to that of NP-sampling. AN-sampling can be performed with less training, reduces patient discomfort, and enables scaling up of antigen testing strategies. Test line intensity however is lower when using AN swabs which could influence negatively the interpretation of the Ag-RDT results. Additional studies on Ag-RDTs using AN swabs on self-interpretation by a lay person are needed and further education around how to interpret a positive Ag-RDT to the wider community.

## Data Availability

All data produced in the present work are contained in the manuscript

## Acknowledgements

Condor steering group: Dr A. Joy Allen, Dr Julian Braybrook, Professor Peter Buckle, Ms Eloise Cook, Professor Paul Dark, Dr Kerrie Davis, Dr Gail Hayward, Professor Adam Gordon, Ms Anna Halstead, Dr Charlotte Harden, Dr Colette Inkson, Ms Naoko Jones, Dr William Jones, Professor Dan Lasserson, Dr Joseph Lee, Dr Clare Lendrem, Dr Andrew Lewington, Mx Mary Logan, Dr Massimo Micocci, Dr Brian Nicholson, Professor Rafael Perera-Salazar, Mr Graham Prestwich, Dr D. Ashley Price, Dr Charles Reynard, Dr Beverley Riley, Professor John Simpson, Dr Valerie Tate, Dr Philip Turner, Professor Mark Wilcox, Dr Melody Zhifang.

We would like to acknowledge the participants for volunteering for this study and to the UK National Institute for Health Research Clinical Research Network and the COvid-19 National DiagnOstic Research & evaluation (CONDOR) programme. We also thank the CRN for their support during the study and Daisy Bengey, Rachel Watkins, and Lorna Finch for their help on sample collection and recruitment.

